# SARS-CoV-2 infection in London, England: Impact of lockdown on community point-prevalence, March-May 2020

**DOI:** 10.1101/2020.05.21.20109017

**Authors:** Michael Edelstein, Chinelo Obi, Meera Chand, Susan Hopkins, Kevin Brown, Mary Ramsay

## Abstract

COVID-19 point prevalence PCR community testing allows disease burden estimation. In a sample of London residents, point prevalence decreased from 2.2% (95%CI 1.4;3.5) in early April (reflecting infection around lockdown implementation) to 0.2% (95%CI 0.03-1.6) in early May (reflecting infection 3-5 weeks into lockdown). Extrapolation from reports of confirmed cases suggest that 5-7.6% of total infections were confirmed by testing during this period. These data complement seroprevalence surveys improving the understanding of transmission in London.

## Background

The World Health Organization declared COVID-19, a respiratory disease caused by SARS-CoV-2, a pandemic on March 11^th^ 2020. The UK reported its first case on January 31^st^; and as of May 18^th^ 2020 243,695 confirmed cases had been reported[1]. At the beginning of the study in early April, UK policy was to prioritise available tests for patients requiring hospitalisation and outbreaks in institutional settings. Evidence suggests that over 80% of COVID-19 cases are mild[2] and that the proportion who are asymptomatic when tested may be as high as 80%[3]. Therefore, using laboratory confirmation in hospitalised cases will severely underestimate disease incidence. Repeated testing among individuals in the community will help estimate trends in COVID-19 point prevalence and help to describe the characteristics of milder infection. We assessed COVID-19 point prevalence in London between early April and early May 2020, which approximately reflect infection around the time of the lockdown and 3-5 weeks into lockdown.

## Use of a participative cohort

Flusurvey (Flusurvey.net) is a participative surveillance cohort set up for influenza surveillance[4] and adapted for COVID19. Any member of the UK public can register and report on symptoms weekly. On March 27th 2020, all Flusurvey participants resident in London were asked for their consent and the consent of their household members, to receive a nasal swab for COVID-19 testing. Self-swabbing has previously been shown to be acceptable to Flusurvey participants[5] and uses packaging compliant with UN3733[6]. Together with the swab, Flusurvey participants and their household members were asked to report any cough, fever, shortness of breath and loss of smell in the two weeks prior to the swab using a short questionnaire. The first set of swabs and questionnaires were posted to participants on 31/03/2020 (wave 1). In the second phase of swabbing, half of the participants were randomly selected and sent a swab between 29 and 30/04/2020 (wave 2). In order to ascertain pre-lockdown point prevalence in the first wave of sampling, we divided wave 1 samples between timely (collected before 7/04/2020) and late (collected or received 7-29/04/2020) samples.

Nasal swabs were tested for viral RNA using polymerase chain reaction (PCR) at the Public Health England Respiratory Virus Unit using primers targeted to either the RdRp and/or Orf gene[7,8].

The age and sex distribution of the initial respondents were compared with the London population[9], and positivity rate by sex and age group calculated, together with 95% confidence intervals (95%CIs). We described the symptoms that timely wave 1 participants reported and their association with being a case, using risk ratios together with 95%CIs. We calculated the positive predictive value of each of the reported symptoms as well as for fever and cough as a cluster. We calculated the positivity rate among participants who returned their first swab late and their second swab, with 95%CIs.

## Characteristics of participants

1,373 individuals (582 London Flusurvey participants and 791 household members) consented to self-swabbing. By 12/05/2020, 1081 first wave samples were received (response rate: 79%) of which 17 were excluded because the participant no longer reported a London address. The participants who sent the remaining 1064 samples were distributed across each London Borough, although the sample underrepresented children (table 1). Age distribution of participants did not differ significantly by sex (p=0.8). Of the 698 individuals who received a second swab, 444 had returned them by 12/05/2020 (response rate 63.6%). Of the 1064 wave 1 samples received, 816 were timely and 248 were late.

**Table 1.** Age distribution of sample compared with London population

| Age group | London |  | Flusurvey participants |  |
| --- | --- | --- | --- | --- |
|  | n | % | n | % |
| <10 | 1215736 | 13.7 | 59 | 6.7% |
| 10-19 | 988894 | 11.1 | 59 | 7% |
| 20-44 | 3626354 | 40.7 | 345 | 43.2% |
| 45-54 | 1160346 | 13 | 119 | 15.3% |
| 55-64 | 857538 | 9.6 | 126 | 14.2% |
| 65+ | 1059213 | 11.9 | 108 | 13.5% |

Of the 816 timely samples from the first wave, 18 were positive in 14 households (positivity 2.2%, 95%CI 1.4-3.5). Positivity was higher among females and among 10-19 year olds (Table 2). Of the 18 positive individuals, 14 (78%) had experienced symptoms in the last 2 weeks and 4 (22%) had not. Symptom onset among symptomatic participants ranged from March 15-30^th^ and time from symptom onset to sample collection ranged from 4-19 days. Cough was the most commonly reported symptom among cases and all symptoms were significantly associated with being COVID19 positive (table 3). The positive predictive value of these symptoms ranged 7.1-13.5% (table 3). Of the 248 late wave 1 samples, 2 (both female and symptomatic) were positive (0.8%, 95% CI 0.2-3.2) and only 1/444 samples received from wave 2 was positive (0.2%, 95% CI 0.03-1.6). The case was female, asymptomatic, not a household contact of a previously identified case, and had tested negative in early April. Of the 20 positive participants in phase 1 of swabbing, five (including three asymptomatic) were retested and all were negative. Of the 3 asymptomatic cases that were retested, none reported symptoms when asked again. Thirty-seven individuals lived in the 16 households which had a least one case. Household size ranged from 1-4 individuals. Five households had 2 cases (Table 4). No additional cases were found during wave 2 in the 5 households that were tested in both waves and had an individual who tested positive in wave 1.

**Table 2.** COVID19 Positivity rate by testing period, age and sex

|  | Participants | Sex |  | Age |  |  |  |  |  |
| --- | --- | --- | --- | --- | --- | --- | --- | --- | --- |
|  |  | Male | Female | <10 | 10-19 | 20-44 | 45-54 | 55-64 | 65+ |
| Testing early April 2020 | COVID19 positive (n) | 5 | 13 | 1 | 3 | 9 | 1 | 3 | 1 |
|  | COVID19 negative (n) | 395 | 403 | 58 | 56 | 336 | 118 | 123 | 107 |
|  | Positivity (%<br>95%CI) | 1.2<br>(0.4;2.9) | 3.1<br>(1.7;5.3) | 1.7<br>(0.04;9.1) | 5.1<br>(1.1;14.1) | 2.6<br>(1.2;4.9) | 0.8<br>(0.02;4.6) | 2.4<br>(0.5;6.8) | 0.9<br>(0.02;5) |
| Testing mid-April 2020 | COVID19 positive (n) | 0 | 2 | 0 | 0 | 1 | 0 | 0 | 1 |
|  | COVID19 negative (n) | 106 | 140 | 12 | 16 | 114 | 44 | 25 | 35 |
|  | Positivity (%<br>95%CI) | - | 1.4<br>(0.3; 5.5) | - | - | 0.9<br>(0.1; 6) | - | - | 2.8<br>(0.4;18.3) |
| Testing Early May 2020 | COVID19 positive (n) | 0 | 1 | 0 | 0 | 1 | 0 | 0 | 0 |
|  | COVID19 negative (n) | 214 | 229 | 35 | 40 | 165 | 74 | 69 | 60 |
|  | Positivity (%<br>95%CI) | - | 0.4<br>(0.06;3.1) | - | - | 0.6<br>(0.08; 4.2) | - | - | - |

**Table 3.**
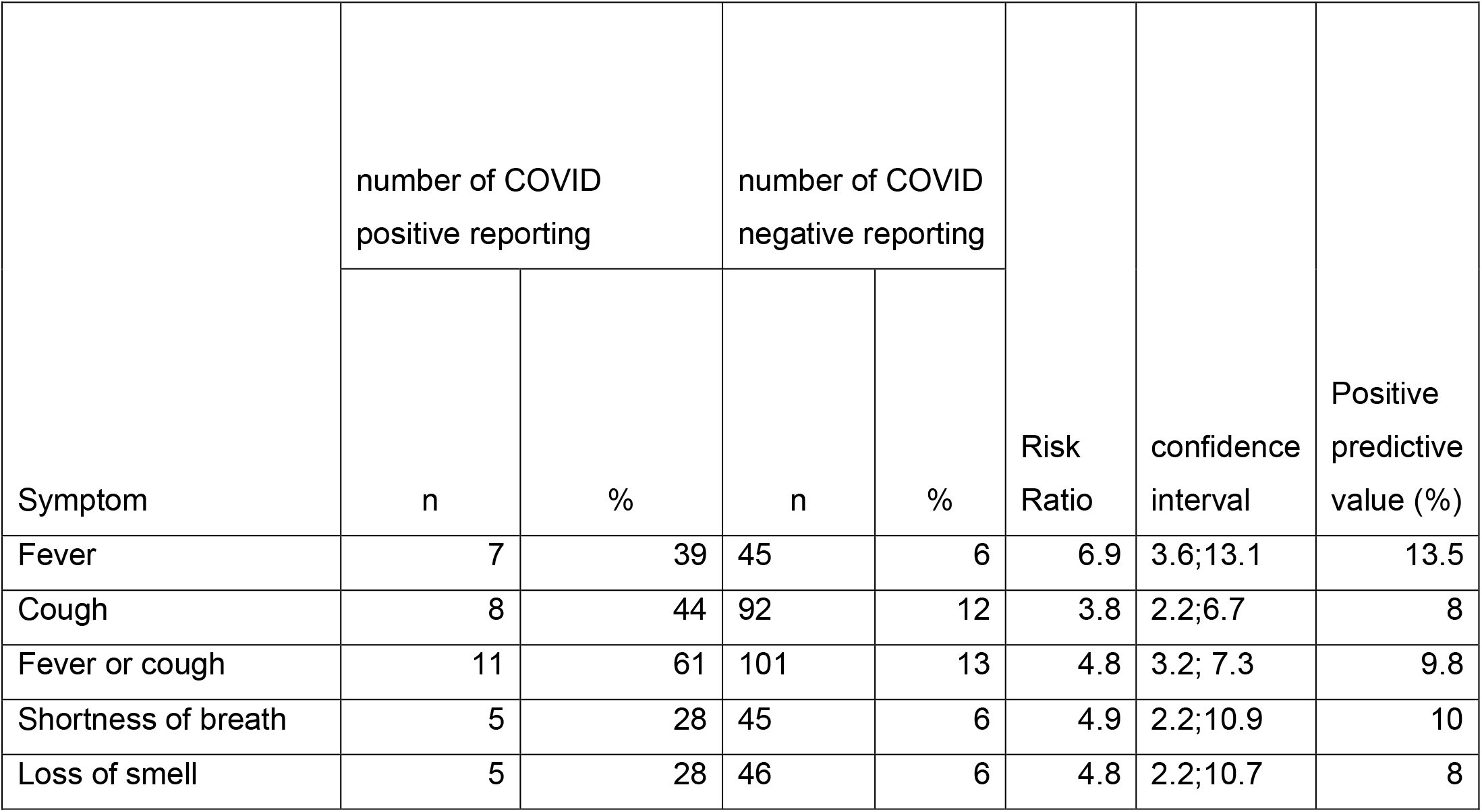
Distribution of symptoms among Flusurvey London participants

| Symptom | number of COVID positive reporting |  | number of COVID negative reporting |  | Risk Ratio | confidence interval | Positive predictive value (%) |
| --- | --- | --- | --- | --- | --- | --- | --- |
|  | n | % | n | % |  |  |  |
| Fever | 7 | 39 | 45 | 6 | 6.9 | 3.6;13.1 | 13.5 |
| Cough | 8 | 44 | 92 | 12 | 3.8 | 2.2;6.7 | 8 |
| Fever or cough | 11 | 61 | 101 | 13 | 4.8 | 3.2; 7.3 | 9.8 |
| Shortness of breath | 5 | 28 | 45 | 6 | 4.9 | 2.2;10.9 | 10 |
| Loss of smell | 5 | 28 | 46 | 6 | 4.8 | 2.2;10.7 | 8 |

**Table 4.** Number of participants positive for SARS-CoV-2 infection among households with at least one participant testing positive.

| **Household number** | **Number of tested household members** | **Number of positives in wave 1 (timely samples)** | **Number of positives in wave 1 (late samples)** | **Number of positives in wave 2** |
| --- | --- | --- | --- | --- |
| 1 | 4 | 1 | - | 0 |
| 2 | 3 | 2 |  | - |
| 3 | 2 | 1 | - | 0 |
| 4 | 1 | 1 | - | 0 |
| 5 | 2 | 1 | 1 | - |
| 6 | 2 | 2 | - | - |
| 7 | 3 | 2 | - | - |
| 8 | 2 | 1 | - | - |
| 9 | 1 | 1 | - | 0 |
| 10 | 2 | 1 | - | 0 |
| 11 | 2 | 1 | - | - |
| 12 | 2 | 1 | - | - |
| 13 | 4 | 1 | - | - |
| 14 | 2 | 2 | - | - |
| 15 | 3 | - | 1 | - |
| 16 | 2 | 0 | - | 1 |

## Impact of the lockdown on point prevalence

This study is the first to describe changes in the point prevalence of COVID-19 infection in the community in London, and England more generally. The 2.2% point prevalence reflects infections acquired in London shortly before and after the lockdown on March 23^rd^ 2020, and the positivity rate of 0.2% reflects infections during the second half of April, three to five weeks into the lockdown. The decrease in incidence suggests the lockdown was effective at reducing COVID-19 transmission in London. Positivity is likely to be an underestimate; apart from the usual limitations of PCR diagnostics, self-testing may be slightly less sensitive than administered swabs[10].

Around the time of our second swab, a pilot survey across England found a similar infection prevalence of 0.27% in early May[11]. In Iceland, a PCR-based screening in a random sample of the population had a 0.6% positivity[12]. In Vo, a town in the Veneto region of Italy, screening of the entire population around the time of the lockdown showed a 2.6% positivity[13]. The London point prevalence in early April was expected to be higher than other UK regions, based on the numbers of hospitalised cases per population at the time of swabbing[1].

## Implications of findings

Evidence suggests that, in mild cases, the virus is detectable in the upper respiratory tract by PCR from approximately 2 days before to 10 days after the onset of symptoms in over 90% of cases[14,15]. A further 10% are positive at 14 days, with occasional reports of longer-term detection[14]. Based on these values, a community PCR point prevalence survey, which only identifies acutely infected individuals, is broadly equivalent to the number of cases acquired over a 12-day period plus an additional 10% of cases acquired in the 4 days before that. Assuming a 5 day lag between disease onset and hospital admission (and testing), and using April 3 as the most common date for the first set of swabs, our infections should correspond to those presenting to hospital and getting tested between March 23 – April 7. Over that time, 10,007 cases were reported in London[1], corresponding to a point prevalence of 0.11%. Compared with the 2.2% positivity from our survey, this suggests that, in London, only 5% of cases were being confirmed through the testing strategy at that time. During the time period around the second swabbing (April 23^rd^ to May 8^th^, determined using a similar approach) there were 2254 reported cases in London, a point prevalence of 0.025% suggesting that 7.6% of infections were detected. These crude figures should be interpreted with caution because viral shedding likely declines over time, affecting test sensitivity in a more complex way. Nevertheless, this figure is consistent with a model that estimated that between 4 and 8.4% of symptomatic cases were being confirmed through testing in hospitals[16].

Two findings were unexpected. First the positivity was higher among females than males, whereas sex-disaggregated data for COVID-19 show equal numbers of cases between men and women[17]. This could be due to a higher proportion of mild cases among females as higher severity among males is well documented[18]. Second, 45% of the cases with a date of symptom onset tested positive more than 10 days after the beginning of their symptoms; this compares published data suggesting 90% of mild cases testing negative 10 days post symptom onset[14]. These findings warrant further investigation.

The repeated questionnaire and swabbing enabled us to confirm that all retested positive cases who reported no symptoms were genuinely asymptomatic rather than pre-symptomatic.

There are limitations to our data. First, the sample is relatively small and purposive (as individuals self-select to register with the cohort) and is therefore likely to not be representative on characteristics beyond age and sex. Secondly, limited information about timing and symptoms makes reconstructing timelines among households difficult. Thirdly, we made a number of assumptions when estimating under-ascertainment, and did not fully take into account the temporal distribution of likelihood of PCR detection. Linking the swabbing results with information collected weekly in Flusurvey may enable us to reconstruct timelines for each case and to document the duration of symptoms and risk factors associated with becoming infected. This data should be interpreted in conjunction with seroprevalence studies, which are ongoing.

## Ethics statement

Ethics approval for inviting participants for self-swabbing was obtained when Flusurvey was initially set up in 2008/9, and each participant was individually re-consented.

## Data Availability

all available data are included in the manuscript

